# Penicillin-clavulanate susceptibility in clinical methicillin-resistant *Staphylococcus aureus* bloodstream infection strains from the CAMERA2 trial

**DOI:** 10.64898/2026.09.20.26363520

**Authors:** N Petersiel, M Duarte-Aguilera, DS Daniel, AS Hayes, JS Davis, BH Howden, SG Giulieri, SYC Tong, Combination Antibiotics for Methicillin-Resistant Staphylococcus aureus (CAMERA2) Study Group

## Abstract

**Objectives:** Among clinical methicillin-resistant *Staphylococcus aureus* (MRSA) bloodstream infection (BSI) strains, we sought to determine the prevalence of β-lactam-β-lactamase inhibitor (BLBLI) susceptibility, and its’ association with oxacillin susceptibility, genotypic factors, and patient characteristics.

**Methods:** Clinical MRSA BSI strains were tested for susceptibility to penicillin-clavulanate by Etest (n=117, 36% of the dataset) and to oxacillin by disk diffusion assay (n=328, 100%). Penicillin-clavulanate susceptibility was defined as MIC≤ 2mg/L and oxacillin disk susceptibility as an inhibition zone >6mm. All strains were whole-genome sequenced, and alleles of *mecA* annotated. The sensitivity and specificity of the oxacillin disk to predict penicillin-clavulanate ‘susceptibility’ were assessed. Clinical and genomic associations with BLBLI ‘susceptibility’ were assessed by calculating odds-ratios (OR).

**Results:** We found 73/117 (62%) and 205/328 (62%) strains to be ‘susceptible’ to penicillin-clavulanate and oxacillin disk, respectively. Oxacillin disk susceptibility and penicillin-clavulanate susceptibility were highly correlated (sensitivity 97.3% and specificity 95.5%). We therefore used oxacillin disk susceptibility as a proxy for penicillin-clavulanate susceptibility. Comparative genomics of the *mecA* gene uncovered 8 alleles. The oxacillin disk ‘susceptible’ phenotype was associated with specific *mecA* alleles, community-associated MRSA sequence-types (OR 72.8, 95%CI 35.4-162), community acquisition of infection (OR 2.09, 95% CI 1.03-4.27), lower Charlson Comorbidity Index (OR 0.90, 95% CI 0.82-0.98) and SOFA (sequential organ failure assessment) scores at presentation (OR 0.88, 95% CI 0.79-0.98).

**Conclusion:** BLBLI ‘susceptibility’ is common among MRSA strains and is strongly associated with the strain genotype. Infections caused by BLBLI ‘susceptible’ strains are associated with specific patients’ characteristics, namely younger age, fewer comorbidities, acquisition in the community and milder presentations.

**Summary:** In an analysis of the CAMERA2 MRSA strains, β-lactam-β-lactamase inhibitors ‘susceptibility’ was strongly associated with the strain genotype and with specific patients’ characteristics, namely younger age, fewer comorbidities, acquisition in the community and milder presentations.

## Introduction

Methicillin resistant *Staphylococcus aureus* (MRSA) is considered inherently resistant to most β-lactams including combinations of β-lactams with β-lactamase inhibitors (BLBLI), because they harbor the *mecA g*ene[1], leading to production of a modified penicillin-binding protein (PBP2a) with low affinity to most β-lactams (except the new generation anti-MRSA cephalosporins, ceftaroline and ceftobiprole).

Shortly after BLBLI combinations were introduced in the 80’s, *in-vivo* studies[2-6], using different animal models, reported their effectiveness in MRSA infections. The premise to these studies was that compared to antistaphylococcal penicillins, penicillin and ampicillin retain relatively better affinity to PBP2a, and together with β-lactam inhibitors that inhibit penicillinase, have the potential to supress cell wall formation. Some of these studies demonstrated eradication of MRSA by BLBLIs from tissues in rat [3, 4] and rabbit[2] endocarditis models, while others showed no benefit, with similar effects to anti-staphylococcal β-lactams[5, 6]. Recently, Harrison *et al*[7] reported that some MRSA strains exhibit low minimum inhibitory concentrations (MIC) to the combination of penicillin-clavulanate. The authors used an epidemiological breakpoint of ≤2 mg/L to define penicillin-clavulanate susceptibility and showed this phenotype could be predicted by the *mecA* allele.

Previous studies described penicillin-clavulanate susceptibility in strains from Europe, America and Asia, without clinical predictors. Our aim was to characterise the penicillin-clavulanate-susceptible phenotype and genotype, and its association with clinical factors among a large collection of MRSA BSI isolates from Australia, New Zealand, Singapore, and Israel, collected as part of the CAMERA2 trial[8].

## Methods

The CAMERA2 trial has been previously published[8]. The study enrolled 352 adult participants (>18) with MRSA BSI in Australia, New Zealand, Singapore, and Israel between 2015-2018.

The trial received ethics approval from the Hunter New England Human Research Ethics Committee (HREC reference number: 15/02/18/3.05). As part of the initial trial, participants provided written informed consent, as described in the primary manuscript by Tong et al. Because this study was a secondary analysis of previously collected data, reconsent of participants was not required.

### Bacterial isolates selection and antimicrobial susceptibility testing

A subset of 117 (36%) previously selected strains[9], representing the most common sequence types collected in the CAMERA2 trial, were tested for susceptibility to penicillin and penicillin-clavulanate by gradient diffusion (penicillin E-test 0.016-256 mg/L, bioMérieux) on Iso-Sensitest Agar (ISA) with or without 15 µg ml^−1^ clavulanic acid, as described by Harrison *et al*[7]. All isolates (n=328) were tested by oxacillin 1µ disk diffusion assay as described by Ba *et al*[10]. We used a “breakpoint” of ≤ 2 mg/L to define penicillin-clavulanate ‘susceptibility’ as previously defined by Harrison et al[7], and >6mm to define oxacillin disk susceptibility.

### Genotype definitions

All isolates have been previously whole-genome sequenced using the Illumina Next Seq[9]. Quality control, assembly of short reads, assembly annotation and multi-locus sequence types (MLST) were performed as described elsewhere[11]. SCC*mec* were determined using sccmec, a tool for typing SCC*mec* cassettes in assemblies (https://github.com/rpetit3/sccmec). Lineages were classified as community-associated MRSA (CA-MRSA) if they were ST1, ST6, ST30, ST45, ST78, ST93, or ST5 carrying SCC*mec* IV or V. Hospital-associated lineages (HA-MRSA) included ST22, ST239, and ST5 carrying SCC*mec* I or II[12].

Mutations in the *mecA* gene and its regulatory region were identified and alleles were annotated as previously described[9] based on the work by Harrison *et al*[7] (Figure 1A). The phylogenetic tree was created as described in [9].

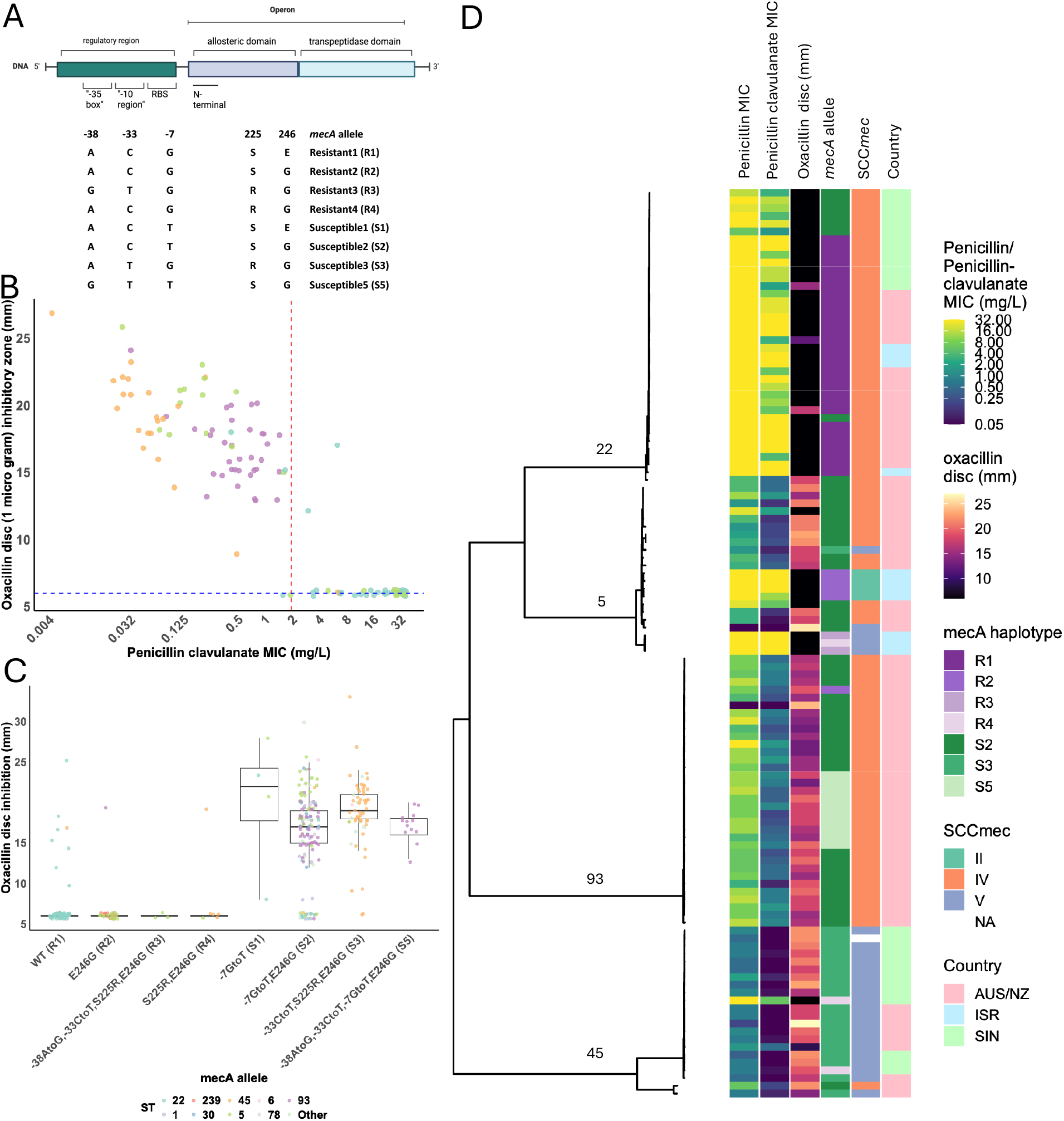

### Statistical analysis

Means are presented with standard deviations and medians with interquartile (25%– 75%) range (IQR). Categorical variables were compared using χ2. Continuous variables were compared by a Student’s t-test if they had a normal distribution or by Mann-Whitney U-test if they had a skewed distribution.

Logistic regression was used to model the association between phenotypic, genotypic and patients’ characteristics and OX disk susceptibility.

Statistical analyses were conducted using R version 2024·09·0.

## Results

Of the 352 participants in CAMERA2, 328 participants had bacterial isolates available for testing. Of these, 117 were tested for both penicillin-clavulanate MIC by Etest and by OX disk diffusion assay to establish oxacillin sensitivity and specificity for determining the penicillin-clavulanate susceptible phenotype.

### Penicillin-clavulanate susceptibility

The median penicillin minimal inhibitory concentration (MIC) for the 117 strains tested was 12 mg/L (IQR 3-32 mg/L), and the median penicillin-clavulanate MIC was 0.75 mg/L (IQR 0.19-16mg/L). The median value and IQR of oxacillin 1µg was 15mm (IQR 6-18) (Figure 1B).

Of the 117 strains that were tested for penicillin-clavulanate susceptibility by Etest and for oxacillin susceptibility by disk diffusion assay, 73 (62%) tested susceptible to penicillin-clavulanate, and 73 (62%) tested susceptible to oxacillin disk (Figure 1B). The sensitivity and specificity of the oxacillin disk assay to determine penicillin-clavulanate susceptibility were 97.3% (71/73) and 95.5% (42/44), respectively.

Because the oxacillin disk showed excellent sensitivity and specificity for predicting penicillin-clavulanate susceptibility and could thus act as an accurate proxy for penicillin-clavulanate susceptibility, we tested the entire CAMERA2 strains cohort (n=328) using the oxacillin disk. Of the 328 strains, 205 (62%) tested susceptible by the oxacillin disk diffusion assay.

### Genomic characteristics of the penicillin-clavulanate phenotype

The most common sequence types (ST) among the 328 strains were ST 22 (n=76), ST 93 (n=57), ST 45 (n=56), ST 5 (n=52), ST 1 (n=20), ST 239 (n=19), ST 30 (n=14), and other STs (n=34) (see phylogenetic tree, Figure 1D).

The rate of phenotypic ‘susceptibility’ by the oxacillin disk differed significantly between the STs (Table 1) and was high for lineages that are considered community-associated (CA) such as ST 93 (56/57, 98%), ST 30 (14/14, 100%), ST 45 (48/56, 86%) and low for lineages that are hospital-associated (HA) such as ST 239 (0/19, 0%) and ST 22 (10/76, 13%). Strains belonging to ST5 exhibited intermediate ‘susceptibility’ (29/52, 56%).

**Table 1:**
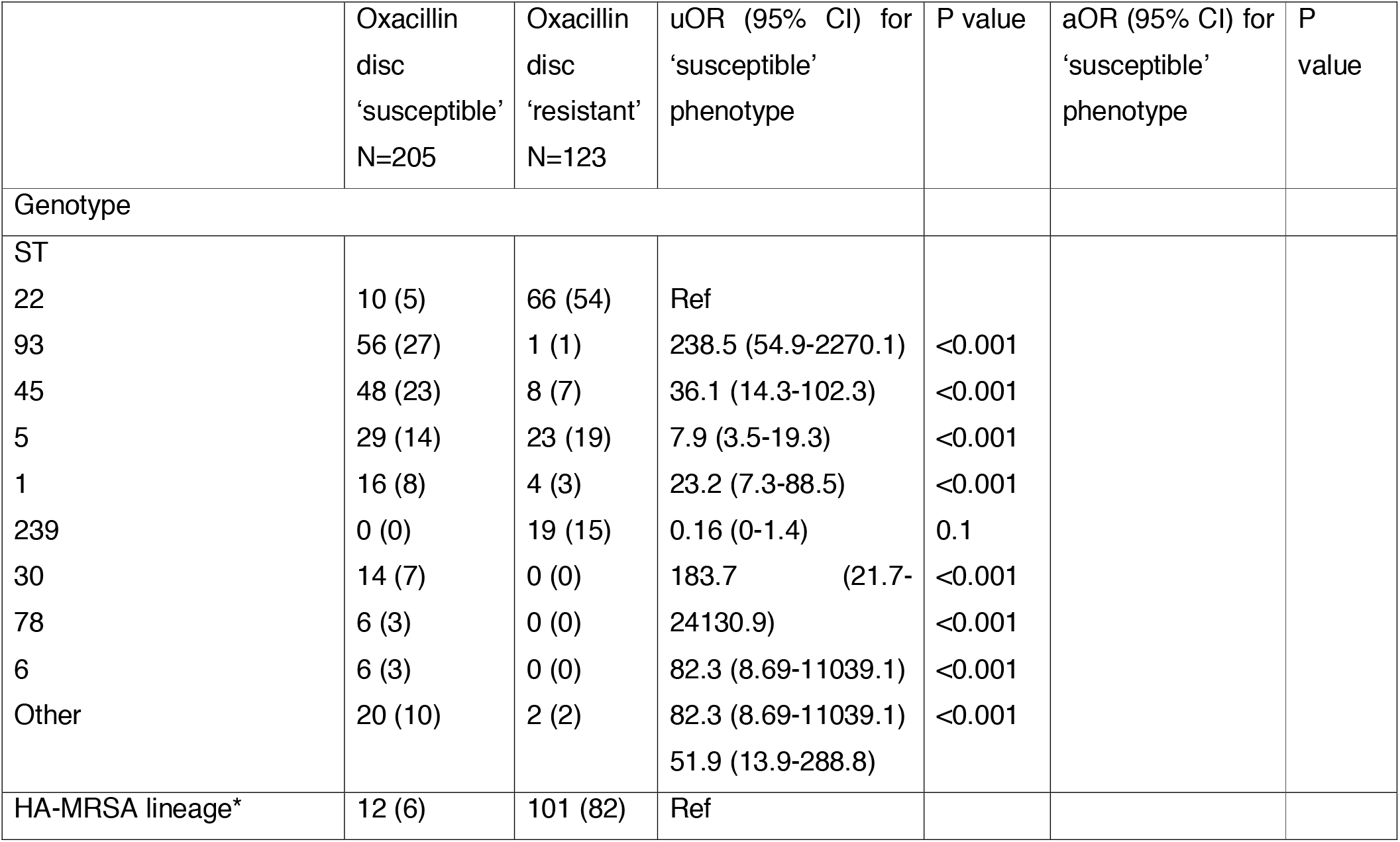
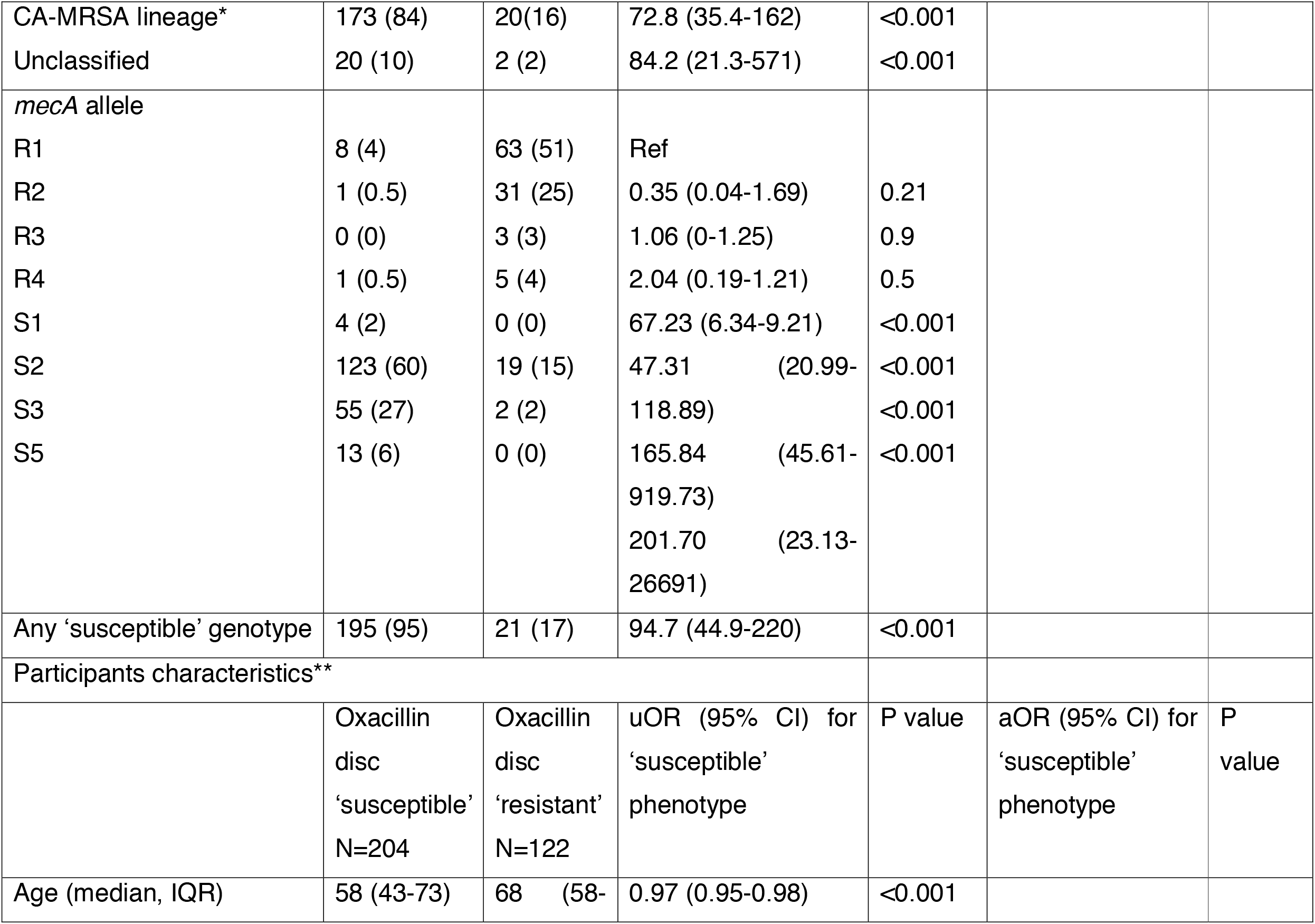
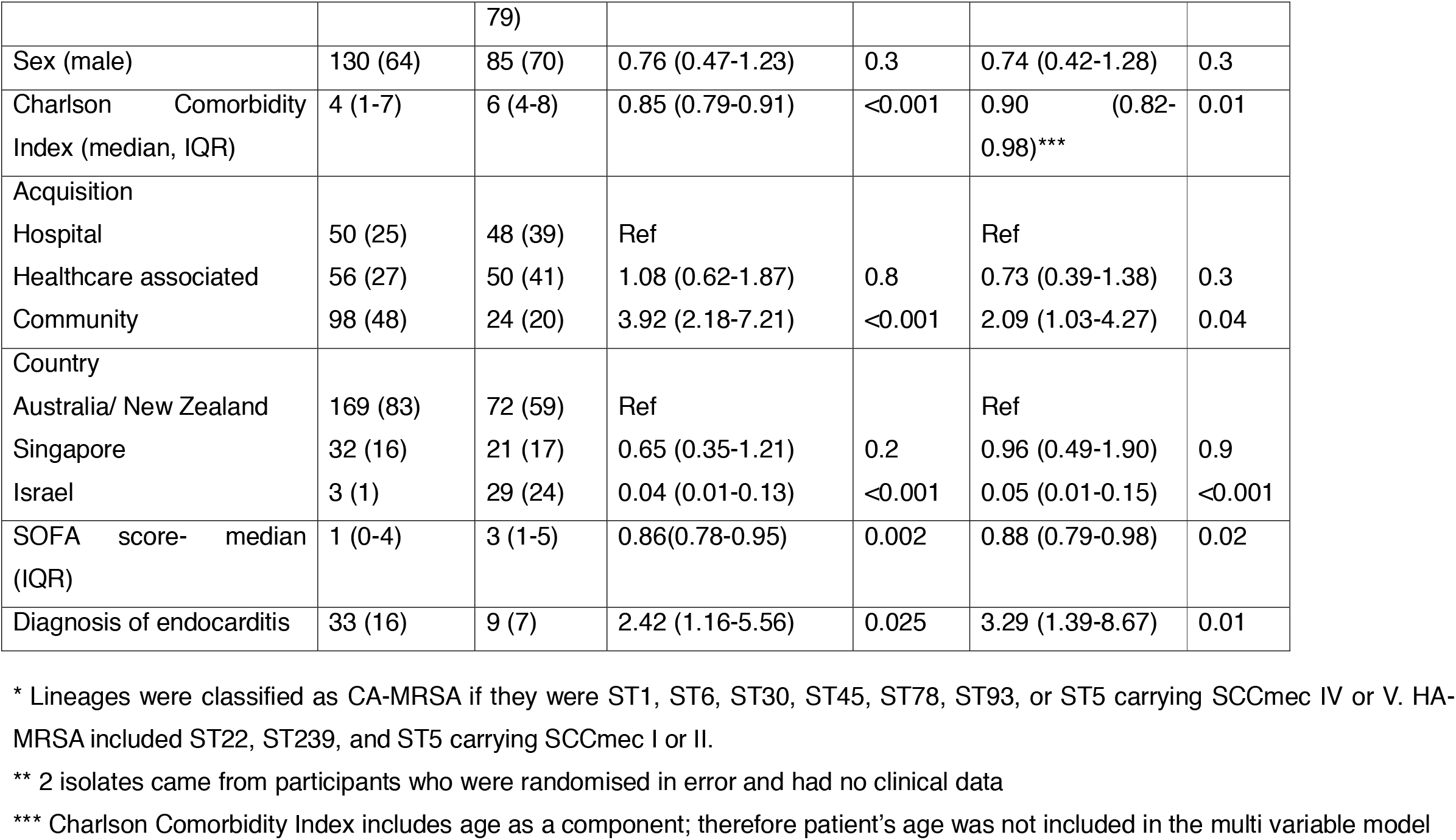
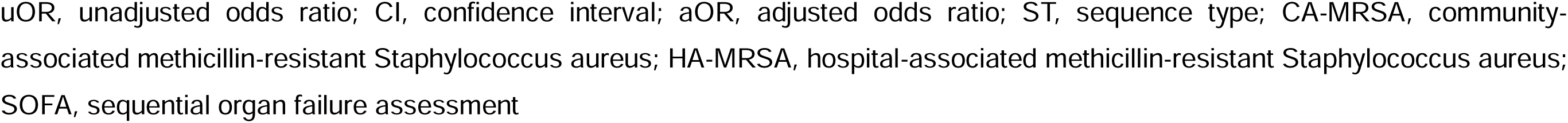
Phenotypic, genotypic and patients’ characteristics and their association with oxacillin dis c susceptibility (defined as inhibition zone>6 mm around 1μg oxacillin disc)

Alignment of the *mecA* gene and its regulatory area (Figure 1A) uncovered 8 *mecA* alleles which were annotated as R1-4 and S1, 2, 3 and 5 based on previous work by Harrison *et a*l[7]. We found 3 new alleles, which we annotated as R3, R4 and S5. Figure 1C shows the distribution of the oxacillin disk inhibition zones (n=328) based on the *mecA* allele. The sensitivity and specificity of the “susceptible” *mecA* genotypes (S1-5) to predict oxacillin disk susceptibility was 95.1% and 82.9%, respectively. The relatively low specificity was driven by 21 false positive results (‘susceptible’ genotype with a ‘resistant’ phenotype), originating from multiple STs, however the majority (19/21) carried an S2 *mecA* allele.

#### Patients’ characteristics

Oxacillin disk ‘susceptibility’ was positively associated with community acquisition (OR 2.09 compared to hospital acquisition, 95% CI 1.03-4.27) and diagnosis of endocarditis (OR 3.29, 95% CI 1.39-8.67), and negatively associated with higher Charlson Comorbidity Index (OR 0.90 for each 1 unit increase, 95% CI 0.82-0.98) and higher SOFA (sequential organ failure assessment) scores (OR 0.88 for each unit increase, 95% CI 0.79-0.98).

## Discussion

In this study of clinical MRSA BSI strains from the CAMERA2 study, the BLBLI ‘susceptible’ phenotype was present in 62% (205/328) of strains and was strongly associated with community-associated clones (ST45, ST93, ST1, ST30), and certain *mecA* alleles (S1-5), compared with BLBLI ‘resistant’ strains. Clinically, the BLBLI ‘susceptible’ phenotype was associated with younger patient age, fewer comorbidities, community-acquired infection, lower SOFA score at presentation and higher rates of endocarditis.

It is important to note that the correlation between the phenotype and the genotype is not perfect. We found 17% (21/123) genetically ‘susceptible’ strains to be phenotypically resistant when tested by oxacillin disk. Similarly, Zhuang et al[13] noted that some isolates show a discordant resistant phenotype with a susceptible genotype and that for these isolates, the *mecA* expression levels were high, similar to isolates with a resistant phenotype and genotype. This emphasises the limitation of using a single genetic factor to predict this phenotype and is an indication that mutations in other genes shape this phenotype[14].

The clinical significance of these findings is uncertain. Previous in-vivo reports have suggested that BLBLI antibiotics may be useful for the treatment of MRSA infections [2-4, 7]. Harrison *et al*[7] demonstrated that BLBLI ‘susceptibility’ predicted MRSA clearance in a murine thigh infection model, where high-dose amoxicillin–clavulanate (100 mg/kg) achieved outcomes similar to vancomycin. However, the doses of amoxicillin-clavulanate used in these studies was higher than the equivalent doses used in humans, and studies using lower doses have shown inferior results compared to vancomycin[5, 6]. Our cohort included too few patients who received a BLBLI (100 participants received either empiric amoxicillin-clavulanate or piperacillin-tazobactam for more than 24 hours pre-randomisation) to warrant a formal analysis of outcomes. Future studies such as the SNAP trial[15] could potentially compare the outcomes of participants with MRSA BSI who received empiric BLBLI according to the infecting isolate BLBLI phenotype.

Our study has limitations. First, our sample size was relatively small. We therefore could not conduct an in-depth genomic analysis of cases where the phenotype and genotype were discordant. Second, as noted above, the study was underpowered to conduct an analysis of the clinical outcomes of patients with MRSA BSI treated with a BLBLI.

## Conclusion

In conclusion, the BLBLI ‘susceptible’ phenotype was prevalent (62%) among MRSA BSI strains from the CAMERA2 study and correlated with CA-MRSA lineages and certain *mecA* alleles. Patients infected by ‘susceptible’ strains were younger with fewer comorbidities.

## Data Availability

All data produced in the present study are available upon reasonable request to the authors

## Acknowledgments

OpenAI ChatGPT (GPT-5.5 and GPT-5.6 Sol; accessed June-September 2026; chatgpt.com) was used solely for grammatical correction and to improve the clarity of author-written text. All AI-assisted edits were reviewed and approved by the authors.

OpenAI ChatGPT (GPT 5.5 and GPT-5.6 Sol; OpenAI; versions accessed in June to September 2026; https://chatgpt.com

The CAMERA2 study group includes the following: David C. Lye, Dafna Yahav, Archana Sud, J. Owen Robinson, Jane Nelson, Sophia Archuleta, Matthew A. Roberts, Alan Cass, David L. Paterson, Hong Foo, Mical Paul, Stephen D. Guy, Adrian R. Tramontana, Genevieve B. Walls, Stephen McBride, Narin Bak, Niladri Ghosh, Benjamin A. Rogers, Anna P. Ralph, Jane Davies, Patricia E. Ferguson, Ravindra Dotel, Genevieve L. McKew, Timothy J. Gray, Natasha E. Holmes, Simon Smith, Morgyn S. Warner, Shirin Kalimuddin, Barnaby E. Young, Naomi Runnegar, David N. Andresen, Nicholas A. Anagnostou, Sandra A. Johnson, Mark D. Chatfield, Allen C. Cheng, Vance G. Fowler Jr, Benjamin P. Howden, Niamh Meagher, David J. Price, Sebastiaan J. van Hal, Matthew V. N. O Sullivan

## References

1. Georgopapadakou, N.H., S.A. Smith, and D.P. Bonner, Penicillin-binding proteins in a Staphylococcus aureus strain resistant to specific beta-lactam antibiotics. Antimicrob Agents Chemother, 1982. 22(1): p. 172–5.

2. Hirano, L. and A.S. Bayer, Beta-Lactam-beta-lactamase-inhibitor combinations are active in experimental endocarditis caused by beta-lactamase-producing oxacillin-resistant staphylococci. Antimicrob Agents Chemother, 1991. 35(4): p. 685–90.

3. Cantoni, L., et al., Comparative efficacy of amoxicillin-clavulanate, cloxacillin, and vancomycin against methicillin-sensitive and methicillin-resistant Staphylococcus aureus endocarditis in rats. J Infect Dis, 1989. 159(5): p. 989–93.

4. Franciolli, M., et al., Beta-lactam resistance mechanisms of methicillin-resistant Staphylococcus aureus. J Infect Dis, 1991. 3(163): p. 514–23.

5. Chambers, H.F., M. Sachdeva, and S. Kennedy, Binding affinity for penicillin-binding protein 2a correlates with in vivo activity of beta-lactam antibiotics against methicillin-resistant Staphylococcus aureus. J Infect Dis, 1990. 162(3): p. 705–10.

6. Chambers, H.F., M. Kartalija, and M. Sande, Ampicillin, sulbactam, and rifampin combination treatment of experimental methicillin-resistant Staphylococcus aureus endocarditis in rabbits. J Infect Dis, 1995. 171(4): p. 897–902.

7. Harrison, E.M., et al., Genomic identification of cryptic susceptibility to penicillins and β-lactamase inhibitors in methicillin-resistant Staphylococcus aureus. Nat Microbiol, 2019. 4(10): p. 1680–1691.

8. Tong, S.Y.C., et al., Effect of Vancomycin or Daptomycin With vs Without an Antistaphylococcal beta-Lactam on Mortality, Bacteremia, Relapse, or Treatment Failure in Patients With MRSA Bacteremia: A Randomized Clinical Trial. JAMA, 2020. 323(6): p. 527–537.

9. Petersiel, N., et al., Genomic investigation and clinical correlates of the in vitro β-lactam: NaHCO(3) responsiveness phenotype among methicillin-resistant Staphylococcus aureus isolates from a randomized clinical trial. Antimicrob Agents Chemother, 2024: p. e0021824.

10. Ba, X., et al., Simultaneously screening for methicillin-resistant Staphylococcus aureus and its susceptibility to potentiated penicillins. J Med Microbiol, 2022. 7(71).

11. Giulieri, S.G., et al., A statistical genomics framework to trace bacterial genomic predictors of clinical outcomes in Staphylococcus aureus bacteremia. Cell Rep, 2023. 42(9): p. 113069.

12. Lakhundi, S. and K. Zhang, Methicillin-Resistant Staphylococcus aureus: Molecular Characterization, Evolution, and Epidemiology. Clin Microbiol Rev, 2018. 31(4).

13. Zhuang, H., et al., A random forest model based on core genome allelic profiles of MRSA for penicillin plus potassium clavulanate susceptibility prediction. Microb Genom, 2021. 7(9).

14. Bilyk, B.L., et al., An Interplay of Multiple Positive and Negative Factors Governs Methicillin Resistance in Staphylococcus aureus. Microbiol Mol Biol Rev, 2022. 86(2): p. e0015921.

15. Tong, S.Y.C., et al., The Staphylococcus aureus Network Adaptive Platform Trial protocol: New tools for an old foe. Clin Infect Dis, 2022.

